# Implementing noncommunicable disease clinics to improve control of hypertension in Somaliland: An observational study

**DOI:** 10.1101/2025.10.26.25338788

**Authors:** Celestin Hategeka, Abdirahman Madar, Yasin Hassan Sh Ali, Safia A. Mohamed, Ahmed Abdi Hirsi, Abdihakim M. Mohamed, Mohamed Ahmed Husein, Max Fraden, Stephen Merjavy

## Abstract

Noncommunicable diseases (NCDs) are increasingly prevalent in Somaliland. However, context-specific evidence on how to successfully implement proven NCD prevention and control interventions in a variety of resource-constrained health systems like Somaliland remains elusive. In 2022, Somaliland’s first pilot NCD clinic, modelled after PEN-Plus in Rwanda, opened at Hargeisa Group Hospital, the national referral and teaching hospital, where patients are treated for heart failure, hypertension, diabetes, COPD, and asthma. The clinic is staffed by nurses and general practitioners with the supervision of an Internal Medicine specialist. The clinic has enrolled over 2000 patients. We evaluate the clinic’s impact on hypertension control to inform improvements and national scale up. A quasi-experimental pre-post study design with mixed effect models was used, analyzing preliminary clinic data from 2022-2024. The outcome was change in systolic blood pressure (SBP) and diastolic blood pressure (DBP) from initial to follow-up visits among hypertensive patients with at least one follow-up. 348 patients (56.6 years and 60% female) were included and had median follow-up of 3 visits (IQR 3) over 114 days (IQR 185). Baseline mean BP was 148.6/96.0 mmHg (95% CI: 139.0–158.3 / 91.9–100.9). Compared with the initial visit, clinic attendance was associated with a significant reduction in SBP ranging from 10.3 mmHg (95% CI: −12.8, −7.7, p<0.0001) at first follow up to 17.3 mmHg (95% CI: −23.4, - 11.1, p<0.0001) at last follow up. Similarly, compared with the initial visit, DBP declined by 5.6 mmHg (95% CI: −7.1, −4.1, p<0.0001) at the first follow up visit and 8.0 mmHg (95% CI: −11.6, −4.3) at the last follow up visit. Implementing NCD clinic was associated with a considerable improvement in hypertension control among NCD clinic patients in Somaliland. Sustaining this reduction in BP would contribute to reducing the risk of major cardiovascular disease events including coronary artery disease, stroke, arrhythmia, and health failure, and ultimately, significant reduction in NCD and all-cause mortality in Somaliland. Lessons learnt while implementing this pilot NCD clinic will help inform successful national scale up of NCD clinic in Somaliland.

## Introduction

Noncommunicable diseases (NCDs), including hypertension, diabetes, heart failure, and chronic respiratory diseases, are an escalating public health challenge in low- and middle-income countries (LMICs).^1–3^ In Somaliland, a state with a fragile health system, NCDs account for over 40% of mortality, with cardiovascular diseases being a leading cause.^4,5^ Despite this burden, resource-constrained settings like Somaliland face significant barriers to NCD prevention and control, including limited healthcare infrastructure, shortages of trained specialists, and high out-of-pocket costs for patients.^4,6^ Context-specific evidence on effective NCD interventions remains scarce, hindering the development of scalable solutions tailored to such environments.^6,7^ As of January 2022, a recent study concluded that none of the hospitals surveyed in Somaliland met the WHO-PEN standard for human resources, equipment, and medicines for effective NCDs management.^8^

To address this gap, Somaliland launched its first pilot NCD clinic in 2022 at Hargeisa Group Hospital, the national referral and teaching hospital, modeled after the PEN-Plus program. PEN-Plus enhances the foundational WHO Package of Essential NCD Interventions (PEN) for decentralization of care for common NCDs including hypertension.^9,10^ First developed and scaled in Rwanda, PEN-Plus has expanded to other LMICs including Haiti, Malawi, and Liberia in partnership with the NCDI Poverty Network.^9,11,12^ This model leverages task-shifting and decentralized care, strategies shown to be cost-effective in other LMICs for addressing NCDs; however, challenges remain in implementing and scaling it in new contexts.^9,11–13^ The Lancet Commission on NCDs and Injuries for the Poorest Billion highlighted PEN-Plus as an effective model for integrated team-based care.^4^ In 2022, the WHO Regional Office for Africa officially adopted PEN-Plus strategy for Africa.^11^

Hypertension, a major risk factor for cardiovascular events such as stroke, arrhythmia, ischemic heart disease, and heart failure, as well as chronic kidney disease is a critical target for intervention, affecting a significant proportion of African population.^1^ Effective blood pressure control can substantially reduce NCD-related morbidity and mortality, yet its implementation in resource-limited settings remains challenging.^14^ We aim to evaluate early impact of the pilot NCD clinic on hypertension control, measured as changes in systolic and diastolic blood pressure, to inform improvements and guide the national scale-up of NCD care in Somaliland.

## Methods

### Study Design, Setting and Population

We employed a quasi-experimental study design to assess the effect of the pilot NCD clinic on changes in systolic and diastolic blood pressures. The study analyzed preliminary data from patients diagnosed with hypertension attending the clinic between December 2022 and December 2024. The study was conducted at the NCD clinic of Hargeisa Group Hospital, Somaliland’s largest public hospital. Patients aged ≥18 years diagnosed with hypertension (SBP ≥140 mmHg or DBP ≥90 mmHg) at their initial visit were included.^15^ Exclusion criteria included incomplete blood pressure records or fewer than two clinic visits. The study was approved by the Hargeisa Group Hospital Ethics Committee. Patient data were anonymized to ensure confidentiality.

### Data Collection

Data were extracted from paper medical records, including demographic characteristics (age, sex) and blood pressure measurements at initial and follow-up visits. Blood pressure was measured using calibrated automated sphygmomanometers by trained nurses following standardized protocols. Follow-up visits occurred every 1–3 months, with the seventh visit as most recent visit within the study period.

### Intervention

The NCD clinic, staffed by nurses and general practitioners under the supervision of an internal medicine specialist, provides integrated care for hypertension, diabetes, heart failure, chronic obstructive pulmonary disease (COPD), and asthma, and is based at the national referral hospital, Hargeisa Group Hospital. Clinical guidelines for the aforementioned entities were approved by the Somaliland Ministry of Health Development (MOHD) and based on PEN-Plus clinical guidelines implemented in public hospital-based clinics in Rwanda.^16^ This model leverages task-shifting and decentralized care, strategies shown to be effective in other LMICs for addressing NCDs as discussed earlier. With over 2000 patients enrolled, the clinic offers subsidized medications and regular follow-up to improve disease management and reduce complications. Physician visits were free of charge for the study duration and laboratory, and medicine costs were subsidized per the discretion of hospital leadership and patients’ ability to pay. The Somalilander American Health Association, a US based healthcare NGO, provided Hargeisa Group Hospital with locally sourced medications in agreement with MOHD. The hypertension protocol defined stage 1 hypertension as BP 140-159/90-99, stage 2 hypertension as BP 160-179/100-109, and stage 3 hypertension as BP > or = 180/110. Patients with stage 1 hypertension and no additional cardiac risk factors were generally considered at low immediate risk. Before initiating therapy in this group, general practitioners advised patients on diet and lifestyle modifications and rechecked blood pressure over a subsequent visit before starting therapy. Patients with multiple additional cardiac risk factors and confirmed stage 1 hypertension were considered for therapy when enrolling in NCD clinic. A more aggressive goal of 130 / 80 was considered reasonable for patients with co morbid diabetes, heart disease, or chronic kidney disease but otherwise below 140 / 90 was targeted. Patients with confirmed stage 2 and 3 hypertension were considered for therapy and often started on two medications with more frequent follow up. First line therapy was thiazide (HCTZ) or calcium channel blockers (amlodipine), however in pregnancy (methyldopa, amlodipine / nifedipine, hydralazine, atenolol), CKD / ESRD (calcium channel blockers, hydralazine, and / or furosemide), heart failure (BB and ACEi/ARB), and diabetes / proteinuria (consider ACEi / ARB) other medications were considered first line. All records in NCD clinic were paper based due to resource limitation (Appendix 1).

### Covariates

Our covariates included variables that have previously been reported to be associated with risk of hypertension including sex, age, smoking, diet, exercise, and currently taking antihypertensive medications.

### Outcome Measures

The primary outcomes were changes in SBP and DBP between the initial visit and subsequent follow-up visit(s). Hypertension control was defined as SBP <140 mmHg and DBP <90 mmHg at follow-up.^15^

### Statistical Analysis

Descriptive statistics were reported as n (%), mean ± standard deviation (SD), or median [minimum, maximum; 25^th^, 75^th^ percentile], as appropriate, to describe the study sample and proportion of patients achieving hypertension control during the study period. We used mixed effects regression models to estimate the effect of clinic attendance on blood pressure changes, adjusting for age, sex, smoking, diet, exercise, financial support, and use of antihypertensive medications within one month preceding the initial visit. Patients contributed data to the analysis until their last visit, with only those with at least one follow-up visit included. A two-sided P*-*value of <0.05 was considered statistically significant. Analyses were performed using R software version 4.5.1.

## Results

Table 1 outlines the characteristics of our study group, comprising 2000 patients from the NCD clinic, with 681 (34.2%) diagnosed with hypertension. Of those with hypertension, 348 (51.1%) had at least one follow up visit. The average age was 56.6 years (SD 12.5) for the hypertension cohort, compared to 44.4 years (SD 16.6) for those without hypertension. Approximately two-thirds of the patients were women. Most patients received financial support including subsidized medications. Type 2 diabetes mellitus was prevalent in our patients (72.6%), while other conditions including type 1 diabetes mellitus (13.4%), heart failure (2.8%), and asthma/COPD (1.7%) were less common. The median number of follow up visits was 3 (IQR, 3), and the median duration between the initial visit and the final follow up was 114 days (IQR, 185).

**Table 1.** Description of the study sample.

|  | All patients in<br>NCD Clinic<br>(N=2000) | Patients without<br>hypertension<br>(n=1309) | Patients<br>with<br>hypertension<br>(n=681) |
| --- | --- | --- | --- |
| **Age, mean (SD)** | 48.62 (16.41) | 44.4 (16.65) | 56.65 (12.51) |
| **Sex** |  |  |  |
| - Male | 714 (36.2) | 475 (36.7) | 238 (35.0) |
| - Female | 1186 (60.1) | 771 (59.6) | 415 (61.0) |
| **Financial support** |  |  |  |
| - None | 108 (5.5) | 73 (5.6) | 35 (5.1) |
| - Partial | 1734 (87.8) | 1150 (88.9) | 582 (85.6) |
| - Full | 133 (6.7) | 70 (5.4) | 63 (9.3) |
| **Tobacco use** |  |  |  |
| - No | 1790 (91.5) | 1159 (90.5) | 629 (93.5) |
| - Current/remote | 166 (8.5) | 122 (9.5) | 44 (6.5) |
| **Qaat\*** |  |  |  |
| - None | 1714 (91.3) | 1111 (89.8) | 601 (94.1) |
| - Remote/current | 162 (8.6) | 124 (10.0) | 38 (5.9) |
| **Exercise<sup>&</sup>** |  |  |  |
| - Inadequate | 1437 (75.6) | 903 (73.0) | 532 (80.4) |
| - Adequate | 462 (24.3) | 332 (26.8) | 130 (19.6) |
| **Diet** |  |  |  |
| - High calorie | 1637 (85.9) | 1055 (84.9) | 580 (87.6) |
| - Healthy | 267 (14.0) | 185 (14.9) | 82 (12.4) |
| **T1DM** |  |  |  |
| - No | 1680 (86.5) | 1041 (80.6) | 637 (98.2) |
| - Yes | 261 (13.4) | 249 (19.3) | 12 (1.8) |
| **T2DM** |  |  |  |
| - No | 480 (24.4) | 290 (22.5) | 189 (27.9) |
| - Yes | 1430 (72.6) | 988 (76.5) | 441 (65.0) |
| **Hypertension** |  |  |  |
| - No | 1309 (65.7) | 1309 (100.0) | 0 (0.0) |
| - Yes | 681 (34.2) | 0 (0.0) | 681 (100.0) |
| **Heart failure** |  |  |  |
| - No | 1936 (97.2) | 1277 (97.6) | 657 (96.5) |
| - yes | 56 (2.8) | 32 (2.4) | 24 (3.5) |
| **COPD** |  |  |  |
| - No | 1763 (98.7) | 1167 (98.7) | 595 (98.7) |
| - Yes | 24 (1.3) | 15 (1.3) | 8 (1.3) |
| **Asthma** |  |  |  |
| - No | 1984 (99.6) | 1301 (99.5) | 681 (100.0) |
| - Yes | 7 (0.4) | 7 (0.5) | 0 (0.0) |

Please note total is less than 100% because of missing data. 609 (89.6%) patients were on antihypertension medications at least one month prior to establishing care with NCD clinic. COPD, chronic obstructive pulmonary disease; NCD, noncommunicable disease; T1DM, type 1 diabetes mellitus; T2DM, type 2 diabetes mellitus. \*Qaat is associated with cardiovascular disease. &Adequate exercise was defined as 30 minutes of activity at least 5 days per week.

At the initial visit, the mean SBP among hypertensive patients with at least one follow-up visit was 148.6 mmHg (95% CI: 139.0, 158.3), and the mean DBP was 96.0 mmHg (95% CI: 91.9, 100.9) (Table 2 and Figures S1 & S2). Compared with the initial visit, clinic attendance was associated with a significant reduction in SBP ranging from 10.3 mmHg (95% CI: −12.8, −7.7, p<0.0001) at first follow up to 17.3 mmHg (95% CI: −23.4, - 11.1, p<0.0001) at last follow up (Table 2, Figure 1). Similarly, compared with the initial visit, DBP declined by 5.6 mmHg (95% CI: −7.1, −4.1, p<0.0001) at the first follow up visit and 8.0 mmHg (95% CI: −11.6, −4.3) at the last follow up visit (Table 2 and Figure 1).

**Figure 1.**
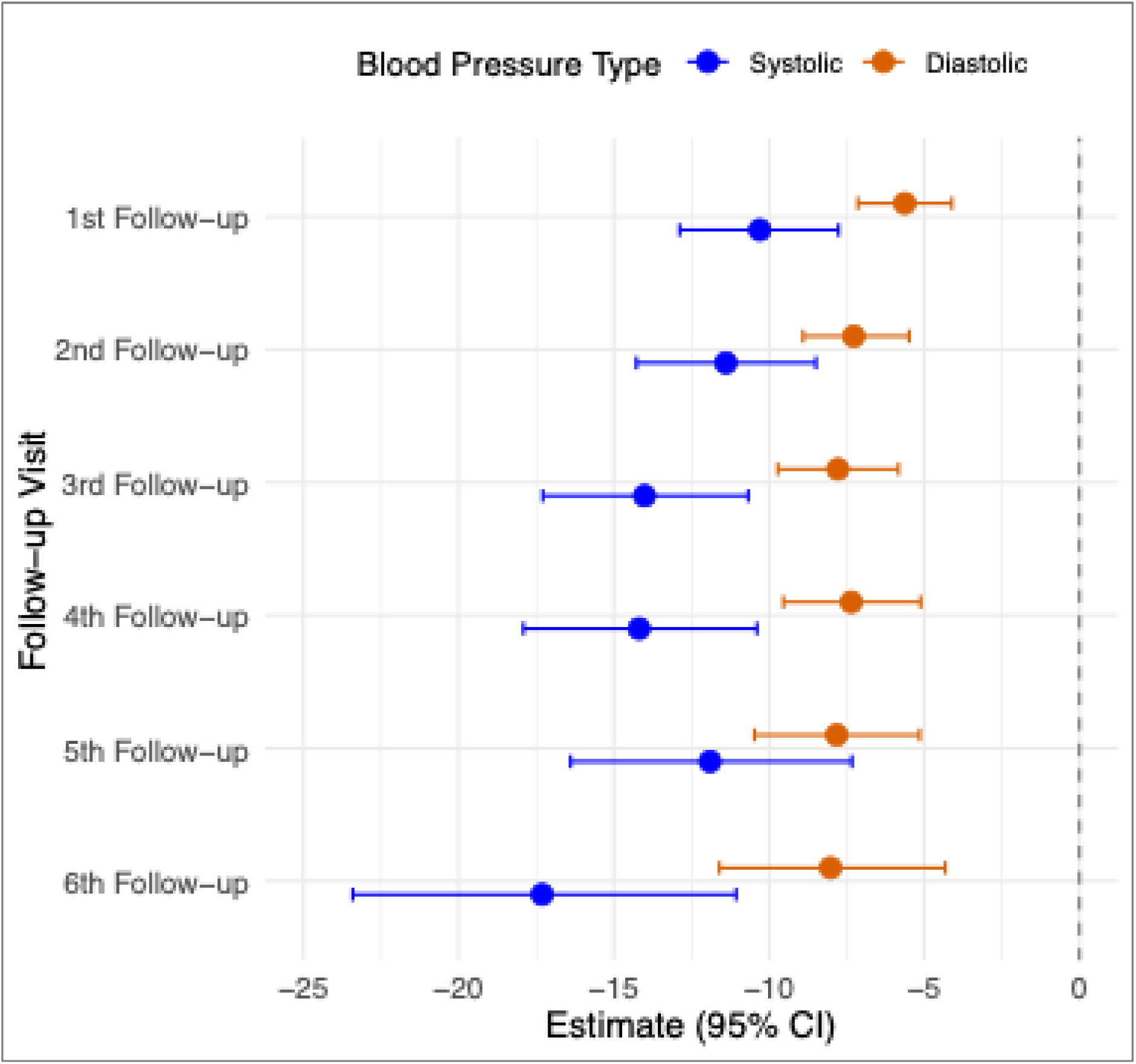
Changes in Blood Pressure with 95% Confidence Intervals. Parameter estimates were derived from mixed effects model assessing association between frequency of follow up visits and changes in systolic and diastolic blood pressures, adjusting for age, sex, exercise, smoking tobacco and Qaat, diet, previously taking antihypertensive medications, and financial support to cover health care cost. These estimates show the patients’ average blood pressure changes at each follow-up visit relative to their baseline. Patients contributed data until their last visit, with only those with at least one follow-up visit included.

**Table 2.** Parameter estimates of association between accessing NCD clinic services and changes in systolic and diastolic blood pressures.

|  | Systolic blood pressure |  | Diastolic blood pressure |  |
| --- | --- | --- | --- | --- |
|  | Estimates<br>(95% CI) | *p* value | Estimates<br>(95% CI) | *p* value |
| Baseline blood pressure | 148.57<br>(139.00, 158.25) | <0.0001 | 96.03<br>(91.07, 100.99) | <0.0001 |
| Change at 1<sup>st</sup> follow up visit | -10.3<br>(-12.88, -7.76) | <0.0001 | -5.63<br>(-7.14, -4.11) | <0.0001 |
| Change at 2<sup>nd</sup> follow up visit | -11.39<br>(-14.27, -8.49) | <0.0001 | -7.27<br>(-8.90, -5.49) | <0.0001 |
| Change at 3<sup>rd</sup> follow up visit | -14.01<br>(-17.29, -10.67) | <0.0001 | -7.78<br>(-9.72, -5.83) | <0.0001 |
| Change at 4<sup>th</sup> follow up visit | -14.18<br>(-17.91, -10.39) | <0.0001 | -7.35<br>(-9.54, -5.12) | <0.0001 |
| Change at 5<sup>th</sup> follow up visit | -11.90<br>(-16.39, -7.33) | <0.0001 | -7.82<br>(-10.47, -5.15) | <0.0001 |
| Change at 6<sup>th</sup> follow up visit | -17.31<br>(-23.41, -11.06) | <0.0001 | -8.02<br>(-11.60, -4.32) | <0.0001 |

Reductions in SBP and DBP were consistent across age and sexes. Overall, 37.1% (range, 33.3-43.1%) of patients achieved hypertension control, and Figure 2 illustrates percentage of patients reaching hypertension control at eat follow-up visit. We noted considerable participant attrition throughout the study, with only 55.2% attending the first follow-up visit, 35.2% at the second, 24.9% at the third, 18.2% at the fourth, 13.1% at the fifth, and 7.0% at the seventh follow-up (Figure 3).

**Figure 2.**
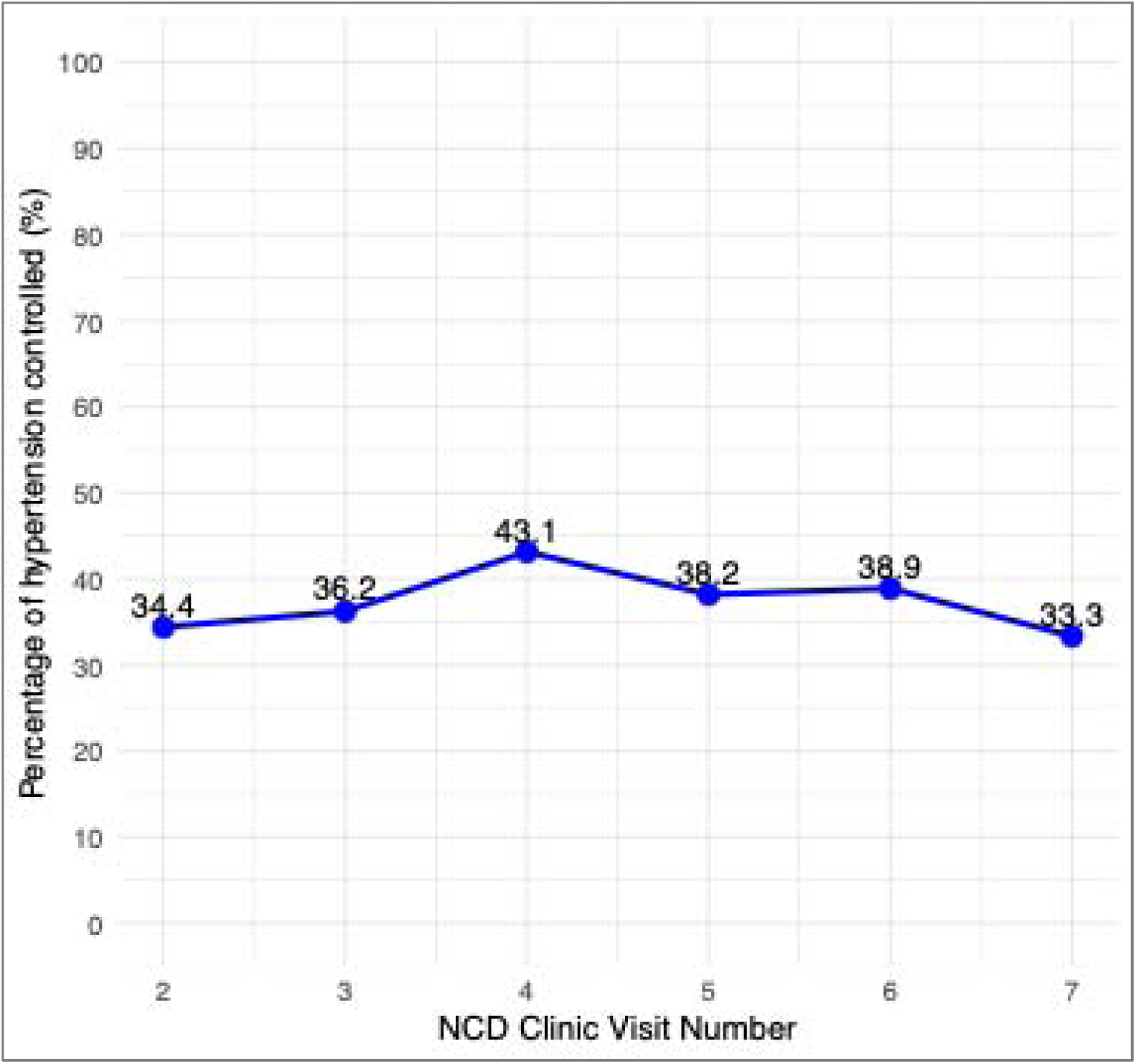
Percentage of patients achieving hypertension control.

**Figure 3.**
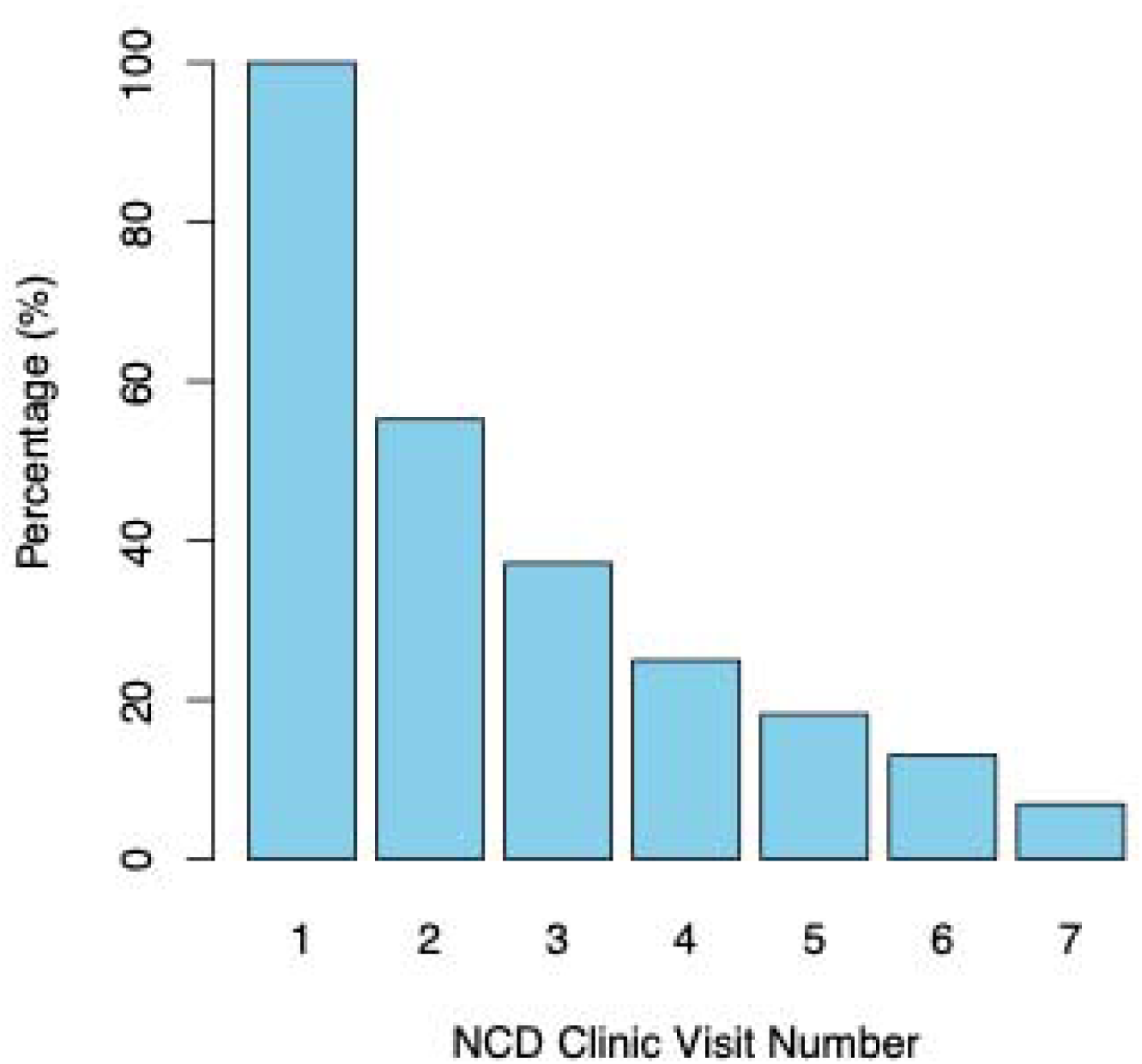
Patients’ attendance rate at each NCD clinic visit.

## Discussion

Our study demonstrates that the pilot NCD clinic at Hargeisa Group Hospital was associated with significant improvement in hypertension control, with mean reductions of up 17 mmHg in SBP and 8 mmHg in DBP among patients with hypertension. These reductions are consistent with prior studies and are clinically significant, as a 10 mmHg decrease in SBP is associated with approximately 20% reduction in major cardiovascular events, including coronary artery disease, stroke, and heart failure.^17,18^ The clinic’s success in achieving these outcomes in a resource-constrained setting like Somaliland highlights the potential of integrated, general practitioner- and nurse-led NCD care models to address the growing NCD burden in low- and middle-income countries (LMICs).^4^

The observed blood pressure reductions align with evaluations of similar decentralized NCD care models, such as the PEN-Plus program in Rwanda, which reported improved outcomes for chronic diseases through task-shifting and integrated care.^12^ The Hargeisa NCD clinic’s reliance on nurses and general practitioners, supervised by an internal medicine specialist, likely contributed to its effectiveness by leveraging existing human resources in a setting with limited specialists. Additionally, the high proportion of patients receiving subsidized medications underscores the importance of affordability in ensuring treatment adherence, a critical barrier in LMICs where out-of-pocket costs often limit access to care.^7^

Despite these promising results, we observed significant loss to follow up during the study period and only 37% of hypertensive patients achieved blood pressure control, suggesting opportunities for further improvement. Patient-level barriers (e.g., health literacy, cultural beliefs around traditional healing, comorbid conditions requiring more urgent care, socioeconomic constraints, travel distance) may have contributed to inconsistent follow-up. For instance, Somaliland’s fragile health system faces challenges like supply chain disruptions and limited diagnostic capacity, which can hinder consistent NCD management in communities closer to patients. Addressing these barriers will be critical for sustaining and enhancing the clinic’s effectiveness.

The quasi-experimental pre-post design, while practical given the nature of the intervention implementation, is a key limitation. The absence of a control group makes it difficult to attribute blood pressure reductions solely to the clinic, as secular trends or patient motivation may have contributed. Additionally, the study relied on preliminary data, and long-term outcomes, such as sustained blood pressure control and reductions in cardiovascular events, remain unknown. Selection bias is another concern, as patients attending the clinic may be more motivated or have better access to care than the broader population with hypertension in Somaliland.

Our findings have important implications for scaling up NCD care in Somaliland. The clinic’s success demonstrates the feasibility of adapting evidence-based models like PEN-Plus to the local context, emphasizing task-shifting, medication subsidies, and integrated care for multiple NCDs (3). Lessons learned include the need for robust supply chains to prevent medication stockouts, ongoing training for non-specialist providers, and community engagement to improve patient retention and adherence. Public-private partnerships and international funding could further support scale-up by addressing resource constraints.

Future research should focus on longitudinal studies to assess the sustainability of blood pressure control and its impact on cardiovascular outcomes in Somaliland. Comparative studies with control groups, including step wedge design, and evaluations of implementation outcomes, such as cost-effectiveness, will also be essential to guide national policy.^6^ Additionally, exploring patient perspectives on barriers to care could inform strategies to improve clinic attendance and treatment adherence. Finally, introducing widespread hypertension screening will detect individuals with hypertension who are unaware of their condition, as seen in many other LMICs.^19^

In conclusion, the pilot NCD clinic at Hargeisa Group Hospital offers a promising model for hypertension management in Somaliland. By addressing implementation challenges and leveraging lessons learned, Somaliland can scale up NCD clinics to reduce the burden of cardiovascular disease and advance progress toward universal health coverage.

## Data Availability

All data produced in the present work are contained in the manuscript.

---

**Appendix 1.**
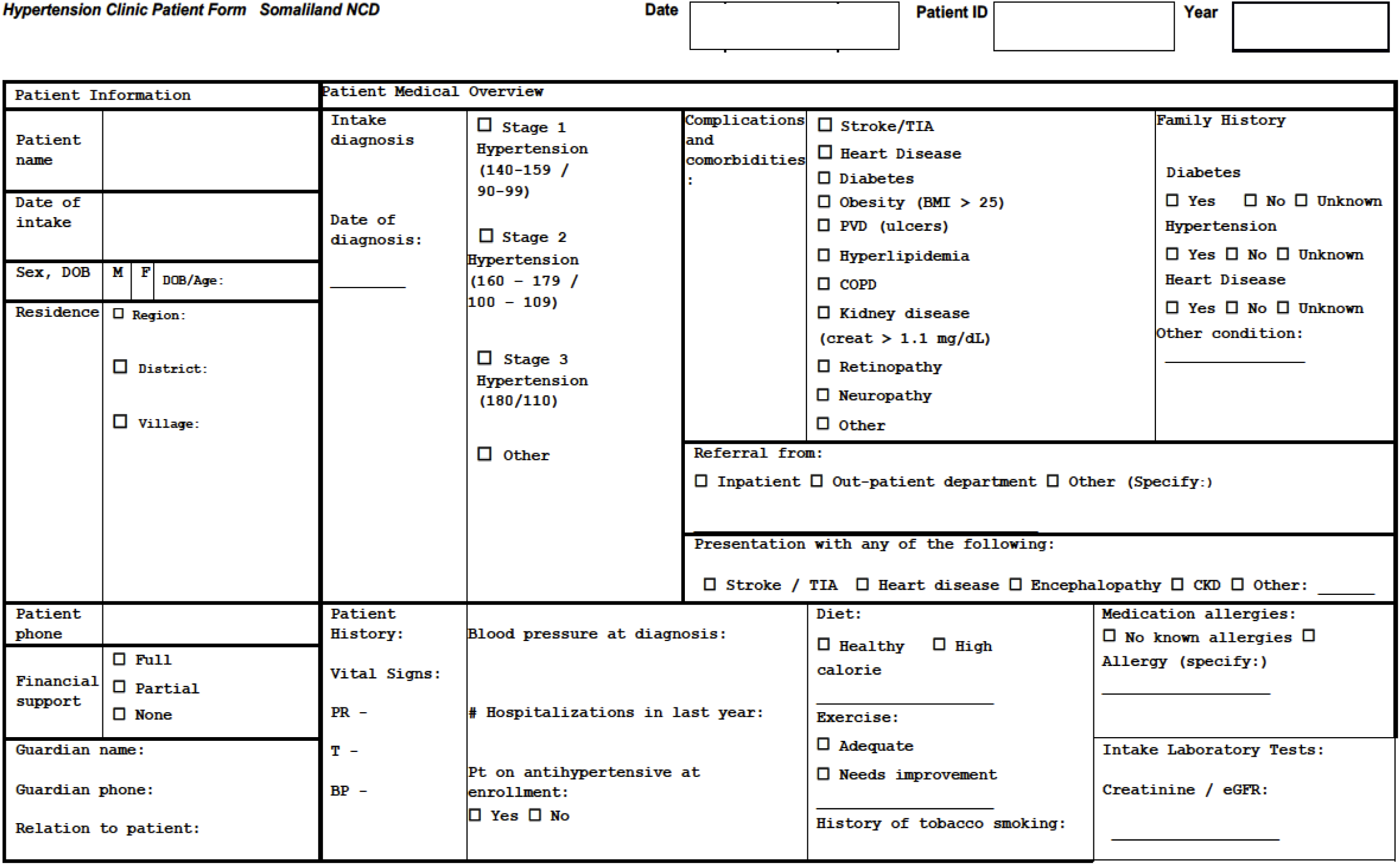
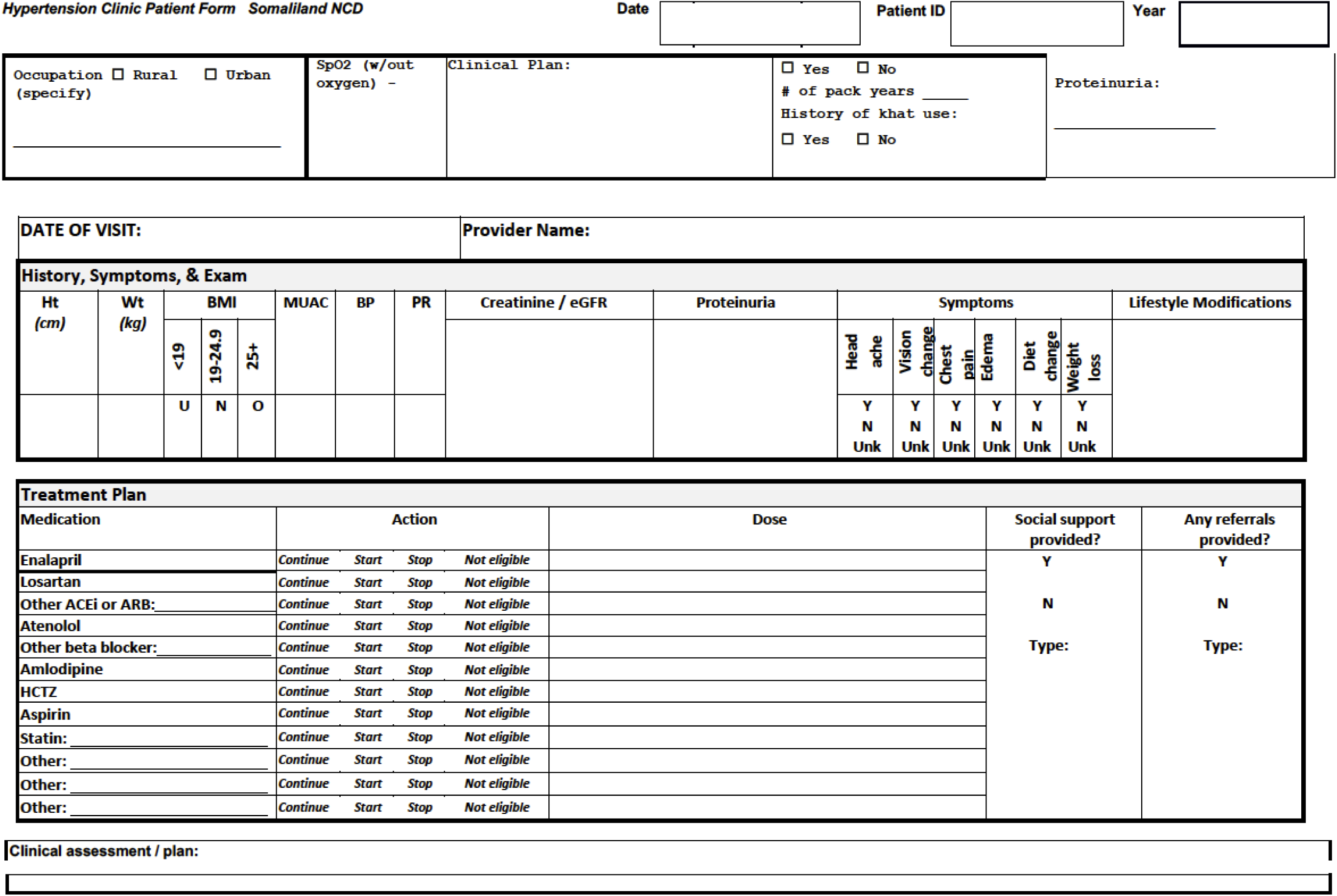
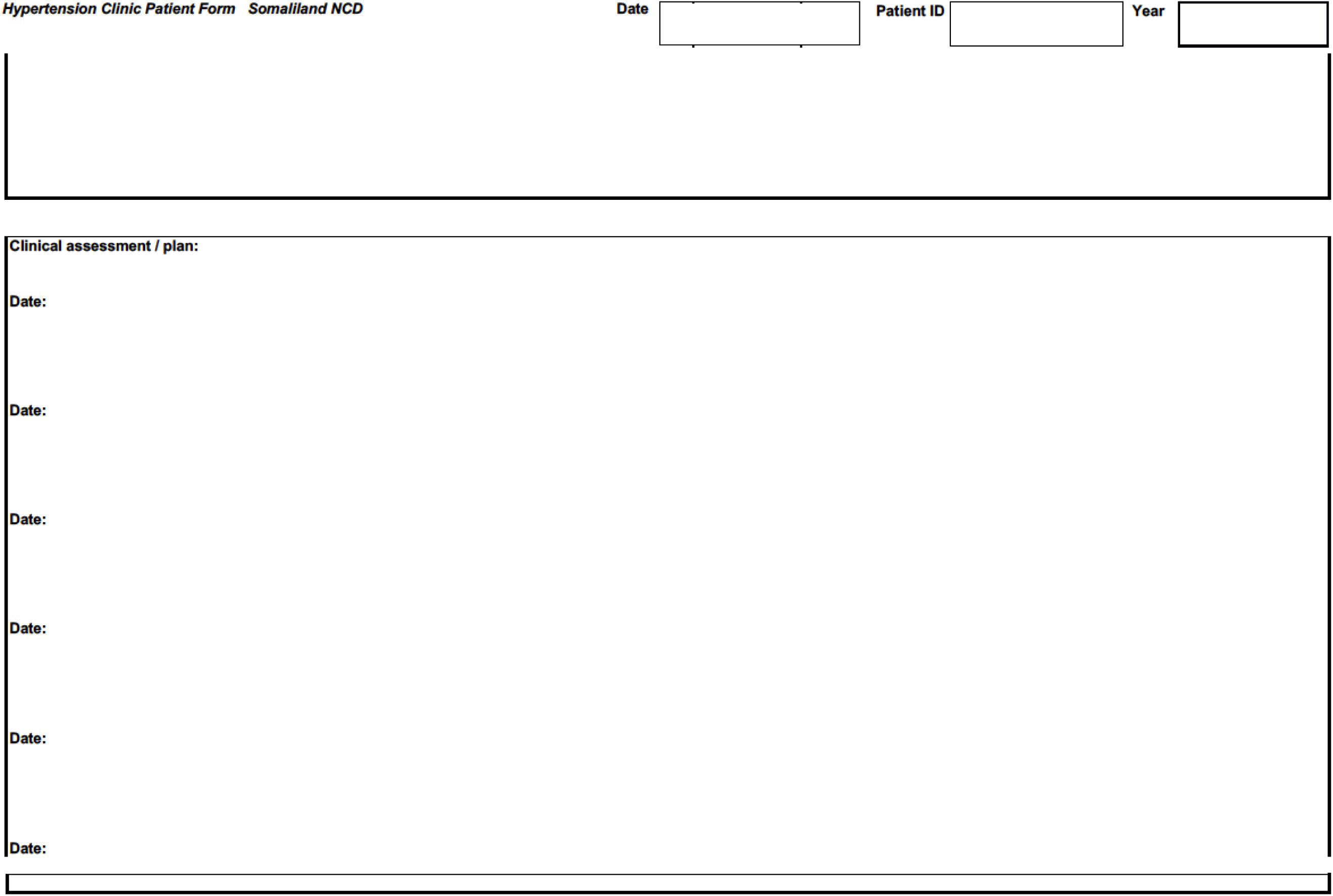
Hypertension Clinic Patient Form Somaliland NCD.

**Figure S1.**
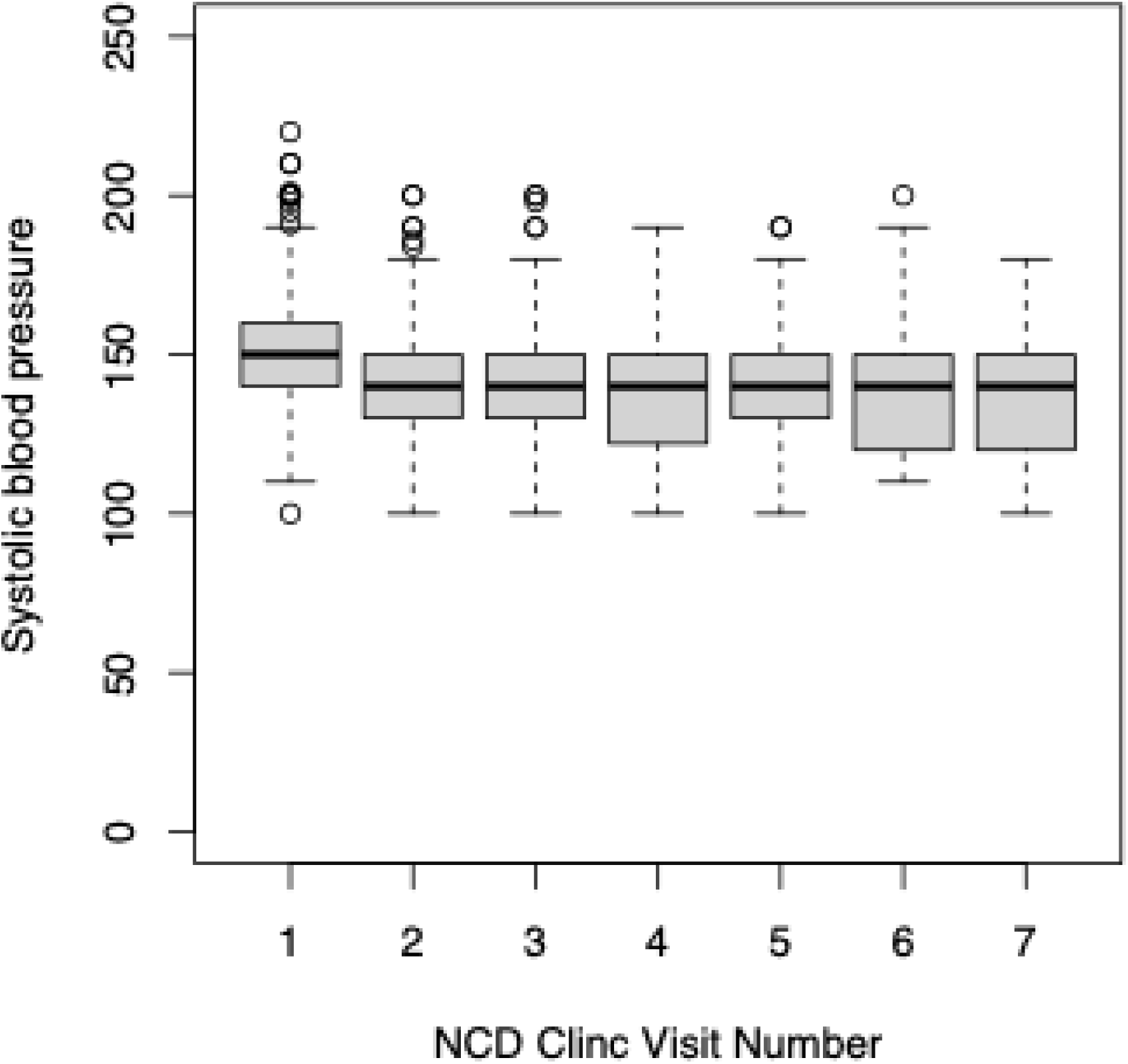
Average systolic blood pressure at each NCD Clinic visit.

**Figure S2.**
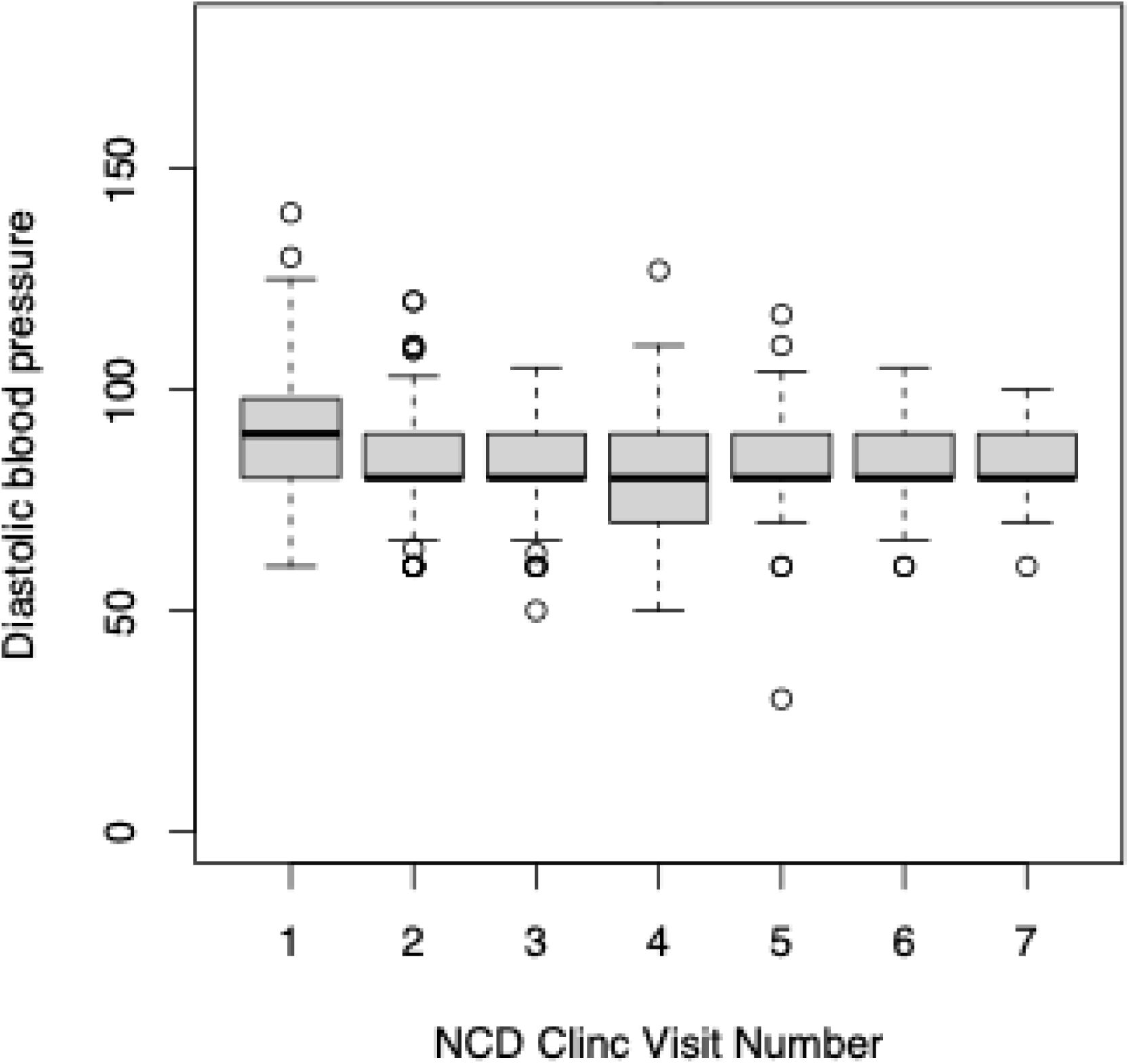
Average diastolic blood pressure at each NCD Clinic visit.

